# County-level Socio-Environmental Factors and Obesity Prevalence in the United States

**DOI:** 10.1101/2023.12.13.23299918

**Authors:** Pedro R.V.O. Salerno, Alice Qian, Weichuan Dong, Salil Deo, Khurram Nasir, Sanjay Rajagopalan, Sadeer Al-Kindi

## Abstract

**Aims:** There are substantial geographical variations in obesity prevalence. Sociodemographic and environmental determinants of health (SEDH), understood as upstream determinants of obesogenic behaviors, may be contributing to this disparity. Thus, we investigated high-risk SEDH potentially associated with adult obesity in American counties using machine learning (ML) techniques.

**Materials and methods:** We performed a cross-sectional analysis of county-level adult obesity prevalence (≥30 kg/m^2^) in the U.S. using data from the Diabetes Surveillance System 2017. We harvested 49 county-level SEDH factors that were used by a Classification and Regression Trees (CART) model to identify county-level clusters. CART was validated using a “hold-out” set of counties and variable importance was evaluated using Random Forest.

**Results:** Overall, we analyzed 2,752 counties in the U.S identifying a national median obesity prevalence of 34.1% (IQR, 30.2, 37.7). CART identified 11 clusters with a 60.8% relative increase in prevalence across the spectrum. Additionally, 7 key SEDH variables were identified by CART to guide the categorization of clusters, including *Physically Inactive* (%), *Diabetes, Severe Housing Problems* (%), *Food Insecurity* (%), *Uninsured* (%), *Population over 65 years* (%), and *Non-Hispanic Black* (%).

**Conclusion:** There is significant county-level geographical variation in obesity prevalence in the United States which can in part be explained by complex SEDH factors. The use of ML techniques to analyze these factors can provide valuable insights into the importance of these upstream determinants of obesity and, therefore, aid in the development of geo-specific strategic interventions and optimize resource allocation to help battle the obesity pandemic.

**Article Highlights:**

- **Why did we undertake this study?** To improve the understanding of the association between complex sociodemographic and environmental determinants of health (SEDH) and obesity prevalence in the U.S.
- **What is the specific question(s) we wanted to answer?** What are the SEDH associated with obesity prevalence?
- **What did we find?** Seven key SEDH variables were identified by CART to guide the categorization of clusters, including *Physically Inactive* (%), *Diabetes, Severe Housing Problems* (%), *Food Insecurity* (%), *Uninsured* (%), *Population over 65 years* (%), and *Non-Hispanic Black* (%).
- **What are the implications of our findings?** Our study shows the importance of SEDH for the regional variation of obesity prevalence and aids in the development of geo-specific strategies to reduce disparities.

## Introduction

The United States is one of the global leaders in the obesity pandemic.^1^ It has been projected that by 2030 almost 1 in 2 Americans will have obesity (body mass index (BMI) ≥ 30 kg/m^2^) and close to 1 in 4 will have severe obesity (BMI ≥ 35 kg/m^2^).^2^ Ultimately, obesity serves as a major risk factor for a plethora of medical conditions such as metabolic disorders (i.e. type 2 diabetes mellitus), cardiovascular diseases, musculoskeletal disease, Alzheimer’s Disease, depression and even some cancer subtypes, making it one of the most important contemporary public health issues.^1^ Although at its core obesity is caused a calorie intake/consumption imbalance, it is crucial to understand its upstream determinants.^3^ While initial focus was drawn to individual-level “obesogenic” behaviors like sedentarism and overconsumption of high-caloric food, more recently, greater attention has been given to the overarching populational-level factors that help shape the downstream effects and influence individual-level obesogenic behaviors.^3,4^

Moreover, these upstream determinants include various sociodemographic and environmental determinants of health (SEDH) that could help understand the substantial county, state and regional-level obesity prevalence variability.^2,5^ However, evaluation of these SEDH variables are often limited by the use of conventional regression-based models, and many are unable to contemplate a broad scope of SEDH factors.^6–8^ Therefore, in this study we set out to explore the relationship of SEDH and adult obesity prevalence across U.S. counties using machine learning (ML) models.

## Methods

### Socioeconomic and environmental data sources (Exposures)

Supplemental Table 1 outlines the 50 variables analyzed in the study which is comprised of county-level SEDH variables that were harvested from previously published sources.^9,10^ The environmental indicators were obtained from the Agency’s Environmental Justice Screening tool (EPA-EJSCREEN) 2020 version (Containing data from 2014 to 2020). These indicators, originally in census block group level, were converted into county-level exposures following the method described in the technical documentation guide of EJSCREEN.^11^ The sociodemographic variables were sourced from the Area Health Resources Files (AHRF) and County Health Rankings & Roadmaps (CHR&R) databases. These SEDH indicators encompass a wide array of fields that include access to healthcare, behavioral risk factors, population characteristics, and other health-related variables.

### Study Endpoints

We analyzed the county-level prevalence of obesity in the United States. Our outcome was defined as the percentage of the adult population (aged ≥18 years) that reports a body mass index ≥30 kg/m^2^ (age-adjusted). This metric was obtained from the Diabetes Surveillance System (DSS) from the Centers for Disease Control (CDC) – 2017. We adopted 2017 data to harmonize with the other 49 variables included in the study. Counties which lacked data were excluded. The descriptive reporting of obesity prevalence was done using the median and interquartile range (IQR).

### Machine Learning and Data Analysis

We employed the Classification and Regression Tree (CART) method. CART is ML technique that was used to delimitate groups of counties that share SEDH characteristics and had similar levels of obesity prevalence (“clusters”). CART is capable of sequentially divide data into smaller, homogeneous groups using binary conditional inferences (“if-then” rules).^12^ To control the splitting of data, we adopted the stopping criteria of a statistical significance (p < 0.05) at each branching point obtained by Pearson’s correlation test, a maximum of depth of 6 splits and a minimum of 150 counties in each terminal node. We then labeled the terminal nodes, or county clusters, with alphabet letters from lowest to highest median obesity prevalence. We validated the CART model using a 20% hold-out sample of the counties.

We evaluated the relative importance of a variable’s association obesity prevalence using Random Forest (RF). RF, another ML technique, aggregates multiple trees using random variable selection and bootstrap sampling.^12^ It then produces an average of the outputs from these trees as a prediction, which we then calculated the relative importance of variables included according to the mean decrease in the node impurity.

We used R v 4.3.0 was used for statistical and ML analyses and QGIS v 3.22.3 for mapping. Statistical significance was determined using a p-value of less than 0.05. No Institutional Review Board approval was needed due to the publicly available nature of the data.

## Results

The study examined 2,752 counties in the United States identifying an overall median obesity prevalence of 34.1% (IQR, 30.2, 37.7). Based on a training dataset comprising 80% of the counties (n = 2204), CART was able to determine 11 terminal nodes (or clusters) of counties that share similar obesity prevalence and SEDH characteristics based on splitting branches (Figure 1). Labeling of these clusters was done in alphabetical order according to their increasing median obesity prevalence (A-K). A 60.8% relative increase in obesity prevalence was observed across the spectrum (from: 25% to 40.2% comparing clusters A and K). To guide county-cluster stratification, CART identified 7 key variables (*Physically Inactive, Diabetes, Severe Housing Problems, Food Insecurity, Uninsured, Population over 65 years and Non-Hispanic Black*) among the 49 potential SEDH to serve as splitting points, each with a statistically significant splitting value. A detailed description of each variable is available in Supplemental Table 1.

**Figure 1.**
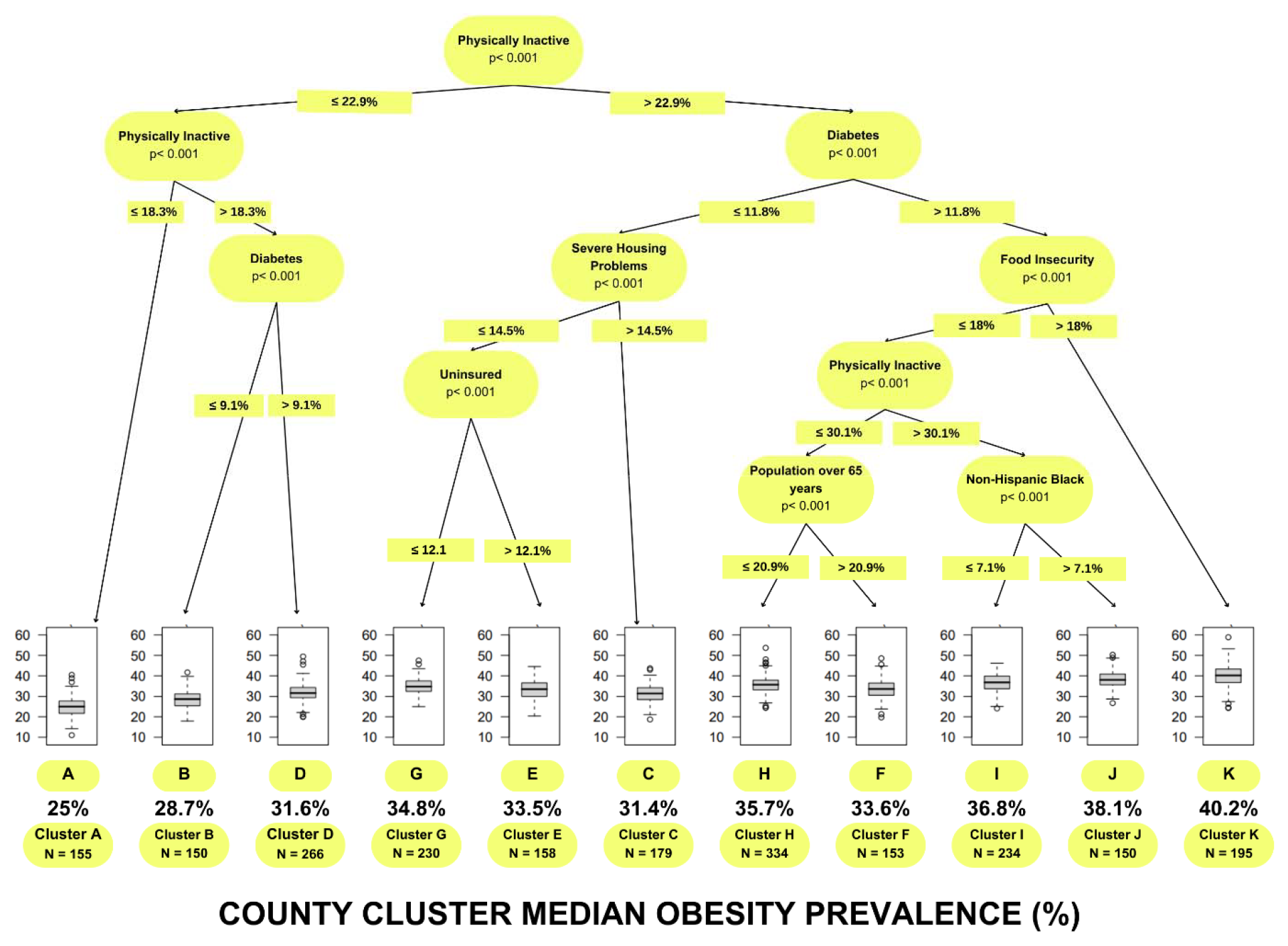
Classification and regression tree (CART) analysis to predict county-level obesity prevalence. Notes: Each path down to a terminal node represents SEDH cluster of counties. Box plots in the terminal nodes represent the median percentage of the adult population (aged ≥18 years) that reports a body mass index ≥ 30 (age-adjusted). The minimum number of counties in a terminal node was set to 150. Clusters were labeled alphabetically based on median obesity prevalence, from lowest to highest.

CART identified *Physically Inactive* to act as the first node, splitting the tree into the left and right side. The left side of the tree (*Physically Inactive* ≤ 22.9%) is comprised of 3 clusters (A, B, D). Furthermore, *Physically Inactive (* ≤ 18.3%) was used again to identify the cluster with the lowest obesity prevalence (Cluster A-25%, IQR: 21.7, 27.7) and separate it from Clusters B and D that had the 2^nd^ and 4^th^ lowest obesity prevalence. The latter differed only in relationship to the percentage of *Diabetes* that was ≤ 9.1% in Cluster B (28.7%, IQR: 25.4, 31.2) and > 9.1% in Cluster D (31.6%, IQR: 29.3, 34.3).

The right side of the tree (*Physically Inactive* > 22.9%) contained 8 clusters (G, E, C, H, F, I, J, K). *Diabetes* was used a second time at the threshold of 11.8% to set apart clusters G, E and C (*Diabetes* ≤ 11.8%) from clusters H, F, I, J, K (*Diabetes* > 11.8%*). Severe Housing Problems > 14*.*5%* was used to delimitate Cluster C (31.4%, IQR: 28.5, 34.2) from Clusters G and E. The latter were differentiated by the percentage of *Uninsured* that was ≤ 12.1% in Cluster G (34.8%, IQR: 32.4, 37.5) and greater than > 12.1% in Cluster E (33.5%, IQR: 30.1, 36.6%). When analyzing the ramifications of *Diabetes* > 11.8% (H, F, I, J, K), *Food Insecurity* >18% could single out Cluster K as the county cluster with the highest obesity prevalence (40.2%, IQR: 36.8, 43.4). Next, *Physically Inactive* was used the third time to separate Clusters H and F (≤ 30.1%) from Clusters I and J (> 30.1%). While Cluster H (35.7%, IQR: 33.2, 37.9%) was characterized by a lower *Population over 65 years* (≤ 20.9%) when compared to Cluster F (33.6%, IQR: 30.5, 36.5), Cluster I (36.8%, IQR: 33.7, 39.9) was characterized by a lower percentage of *Non-Hispanic Black* (≤ 7.1%) when compared to Cluster J (38.1%, IQR: 35.6, 40.1).

Based on the splitting points and the respective threshold values obtained from the regression tree created using the training data, we labeled (Clusters A-K) the remaining 548 counties (test data). We then compared the median obesity prevalence across the training and testing set which showed no substantial differences across the 11 clusters (Supplemental Figure 1). Figure 2A and 2B map the geographic distribution of county-level obesity prevalence and the CART-identified county clusters (A-K), respectively.

**Figure 2.**
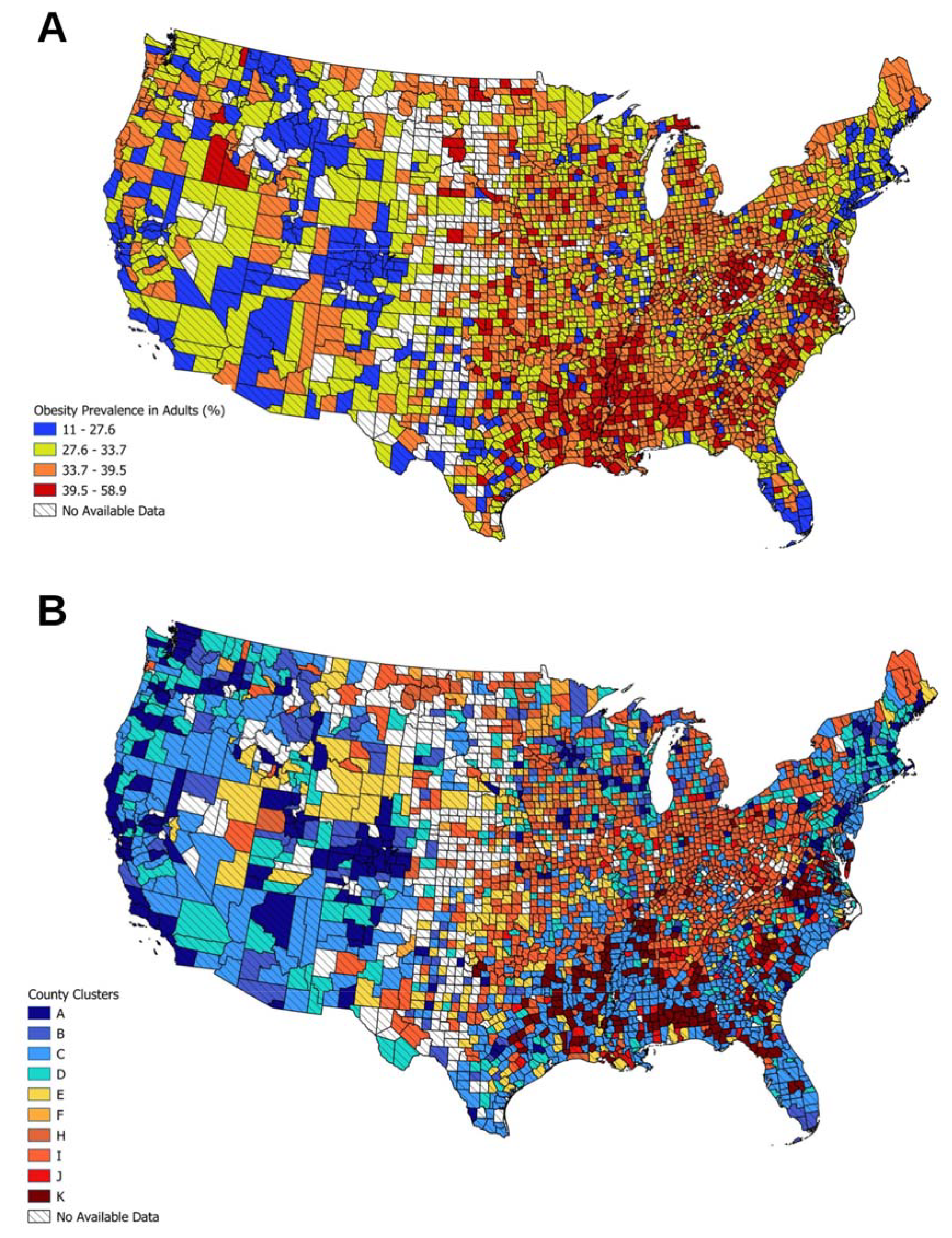
Map of the county-level prevalence of obesity (%) in the United States, divided by natural breaks (A). (B) County clusters identified by CART (A-K), labeled according to lowest to highest obesity prevalence (%).

Finally, we employed RF to assess the relative variable importance in respect to the county-level obesity prevalence and the results are displayed in Figure 3. Overall, from the 49 SEDH variables available, the 7 variables selected by CART were ranked as 1^st^ (*Physically Inactive*), 2^nd^ (*Diabetes*), 7^th^ (*Non-Hispanic Black)*, 8^th^ (*Population over 65 years*), 11^th^ (*Food Insecurity*), 19^th^ (*Severe Housing Problems*), 21^st^ (*Uninsured*).

**Figure 3.**
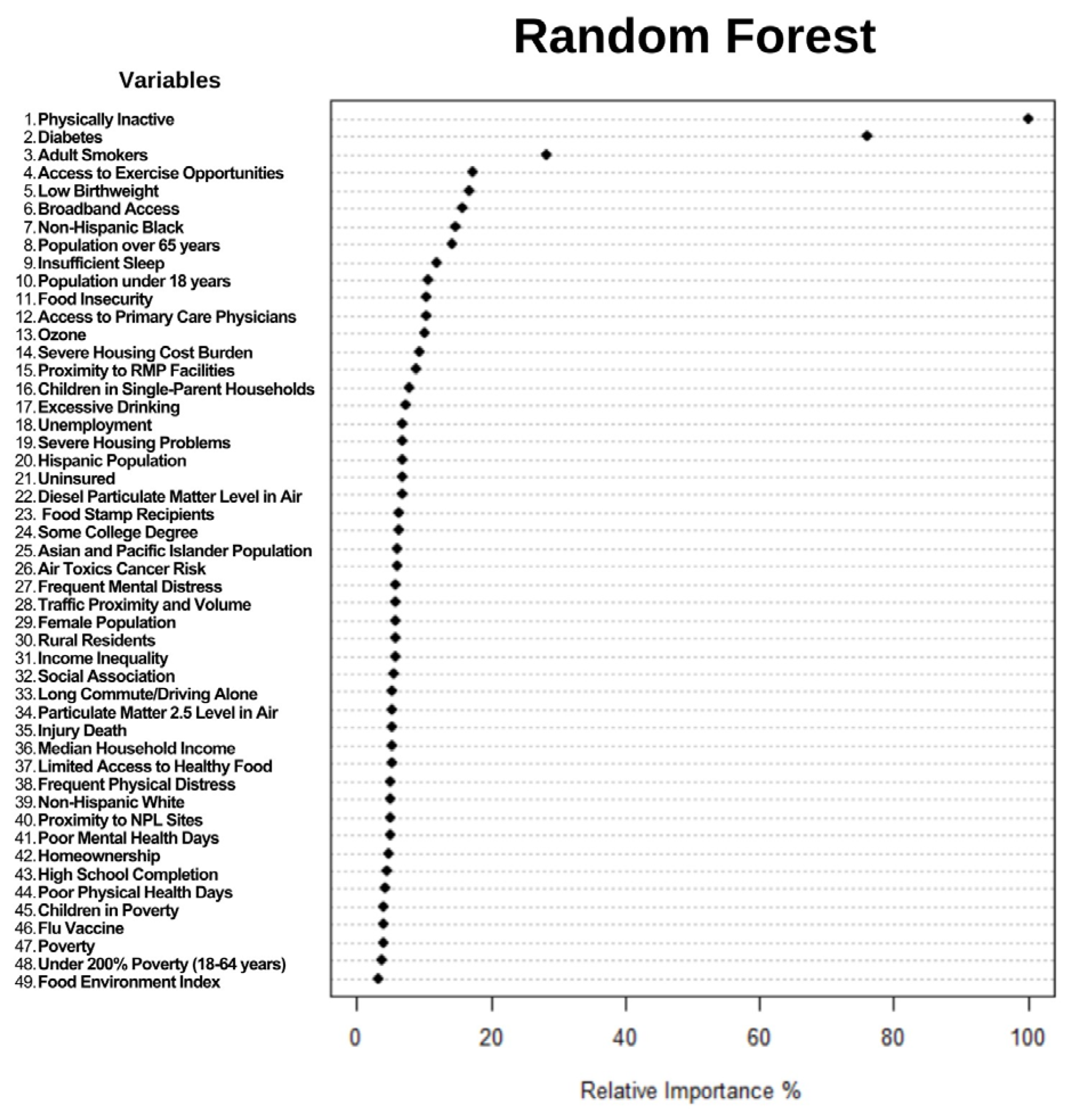
Dot chart of Random Forest analysis displaying variable importance for predicting county-level obesity prevalence in the United States. Notes: the most important variable is at the top and scaled to 100%. The importance of the rest of the variables is shown relative to the top one. Abbreviations: PM, fine particulate matter; RMP, Risk Management Plan; NPL, National Priorities List.

## Discussion

Obesity has been an escalating public health issue and point of discussion for several decades, given its increasing prevalence among Americans. According to the CDC, the age-adjusted prevalence of obesity in U.S. adults was 42.4% in 2017-2018 and continues to rise despite a temporary plateau seen in 2009-2012.^13^ By 2030, most adults are projected to be overweight or have obesity and approximately 50% of adults will have obesity.^14^ Recognizing that obesity contributes to a growing prevalence of chronic disease such as cardiovascular disease, diabetes, gastrointestinal disease and arthritis, and some types of cancer, obesity is regarded as one of the greatest threats to public health of the century.^15,16^ Despite the American Medical Association recognizing obesity as a disease with multiple pathophysiological aspects requiring interventions, little progress has been made in reversing the obesity epidemic.^17,18^ Implementation of prevention strategies have largely focused on risk factors related to food supply, physical activity, and behavior changes, which likely reflect the complexity of the disease that continues to challenge healthcare professionals and researchers.^19^ In this investigation, ML techniques were used to explore the associations between SEDH and adult obesity. CART was subsequently used to provide a clinically applicable visual representation of county-level obesity prevalence. In our assessment, *Physically Inactive, Diabetes, Non-Hispanic Black, Population over 65 years, Food Insecurity, Severe Housing Problems*, and *Uninsured* were identified as associated to obesity prevalence. CART implemented these variables to identify 11 clusters of counties with 60.8% relative increase in prevalence between the lowest and highest risk county clusters. Additionally, Random Forest–an alternative ML approach– illuminated CART’s ability to classify complex SEDH by determining the relative importance of each variable.

CART identified *Physically Inactivity* as the starting node, which was also the most important variable according to Random Forest. “*Physically Inactive”* was utilized twice more as a node further downstream, highlighting the importance of activity as a variable accounting for large variations in obesity prevalence. Studies have shown a positive risk association between physical inactivity as well as sedentary lifestyle with obesity prevalence. Higher rates of sedentary behavior and physical inactivity exist among people with obesity.^20^ Irrespective of BMI, higher physical activity intensity levels and lower sedentary time are associated with lower risk of mortality, and based on these relationships, a combination of lifestyle alterations revolving around being active and reducing time spent sedentary contribute to prevention of abdominal obesity.^21,22^

In addition to Physically Inactivity, *Diabetes* held significance by CART, being utilized twice in the stratification of clusters. This finding coincides with the Random Forest, which identified Diabetes as the 2^nd^ most important variable in predicting obesity prevalence among adults. There are significant associations between obesity and diabetes among other chronic conditions including high blood pressure, cardiovascular disease, high cholesterol, asthma, and arthritis.^15,23^ Likewise, obesity and overweight represent modifiable risk factors for type 2 DM and as many as 80% people are overweight when diagnosed.^15^

Demographic factors seemed to play an important role in predicting obesity prevalence. Specifically, CART identified *Non-Hispanic Black* and *Populations Over 65 Years* as two demographic SEDH variables. Although variations have been reported across studies, obesity prevalence in the United States varies by ethnicity. One study demonstrated that non-Hispanic Black individuals were most likely to have obesity (prevalence of 36.1%) and that individuals living in communities with >25% non-Hispanic Black or Hispanic had significantly higher BMIs and were more likely to have obesity.^24^ Meanwhile, another study showed that highest prevalence of relative fat mass (RFM)-defined obesity was observed in Mexican American men and women and highest prevalence of BMI-defined obesity was highest in African American women.^25^ Previous studies have shown also demonstrated age-related disparities across obesity distribution. While it is unclear whether there is a definite positive correlation with older age and obesity, which can vary depending on the surrogate of body fat percentage, age-related trends in obesity are likely explained by age-related changes in body composition. ^25,26^ After ∼60 years, the proportion intra-abdominal fat progressively increases and energy intake and total energy expenditure declines primarily due to decreases in physical activity and basal metabolic rate. Regardless of age, obesity is still associated with disorders that increase functional.^26^

Like many other public health concerns, socioeconomic status (SES) varies with obesity prevalence. Traditional SES variables identified by CART included *Food Insecurity, Severe Housing Problems, and lack of insurance*. SES affects both the quantity and quality of food consumed. Low-cost foods are highly processed and contain refined grains, sugars, and added fats, which are linked to rising obesity rates. Not only do these food options seem to be of best value by combining low cost with high energy density, but they may also be the most accessible, particularly in “food deserts”. In fact, lower-income households tend to consume more “obesogenic” foods while “protective” foods such as whole grains, lean meats, fish, low-fat dairy products, fresh vegetables, and fruit are more likely to be consumed by groups with higher SES.^27–30^ High housing costs similarly have potential to worsen the obesity epidemic. Populations living in areas of high rent burden are more likely to have obesity, which has been demonstrated by modelling investigations in Chicago, IL. When households spend >50% of their income on housing, access to healthy foods once again becomes a burden.^31,32^ Beyond food and housing insecurity, lack of health insurance represents an additional risk factor associated with adult obesity. Analyses have revealed disparities in obesity prevalence in disadvantaged neighborhoods and conversely, people with obesity constitute one population that are more often uninsured. ^33–35^

The current regime of obesity treatment entails a “treatment pyramid”. Lifestyle interventions serves as the base of the pyramid followed by pharmacological agents, endoscopic procedures, and ultimately bariatric surgeries.^36^ Despite the limitations of both low-to high-intensity lifestyle modifications, the represent the foundation of any weight loss. Several resources such as the USDA and HHS’ *Dietary Guidelines for Americans 2020-2025* and CDC’s *Recommended community strategies and measurement to prevent obesity in the United States* target these potential lifestyle modifications. The former relies on the premise that “no matter their age, race, or ethnicity, economic circumstances, or health status, can benefit from shifting food and beverage choices to better support healthy dietary patterns”.^16^ The latter provides strategies to promote availability of affordable healthy food and beverages, encourage breastfeeding, and support physical activity or limit sedentary activity.^37^ These targeted interventions are represented in this study by *Physical Inactivity* and to some extent, *Food Insecurity*. While lifestyle risk factors are of vital importance, our study demonstrates that other SEDH-related factors can aid in identification of areas with higher obesity prevalence, and therefore, deserve more attention in future investigations. Moreover, use of tree-based ML approaches such as CART provides a practical representation of the intricate web of SEDH and geographic variation in obesity prevalence.^10,12^

This study has limitations. Newer research suggests that BMI is not an adequate surrogate for visceral adiposity. BMI does not distinguish between visceral fat and subcutaneous fat, which varies with age, gender, and ethnicity. For example, women have more subcutaneous fat than men on average and Asians have more visceral adiposity than Caucasians and typically have much smaller BMI’S. BMI is not surprisingly less predictive in many populations with higher visceral adiposity including elderly populations. Direct measurement of visceral adiposity could provide much needed specificity in the obesity and will help clarify historic relationships or lack thereof.^26^ Another limitation is the cross-sectional characteristic of this study which is associative by nature. This study focused only on risk factors represented by the 49 SEDH variables in the model, and therefore other factors such as genetics, family history, and medications were not incorporated. We excluded counties with no data and future methods may consider consolidation. However consolidation together with county level analysis may discount significant within county variation.^39,40^

## Conclusion

The application of machine learning techniques to a nationwide prevalence data identified *Physically Inactivity, Diabetes, Non-Hispanic Black, Population over 65 year, Food Insecurity, Severe Housing Problems*, and *lack of insurance* as key SEDH variables that delineated clusters of U.S. counties sharing similar rates of obesity prevalence. Comprehensive visualization of these clusters using Classification and Regression Tree (CART) demonstrated significant county-level geographic variation of obesity prevalence in the United States, which provides the potential to advance public health initiatives and policies surrounding obesity.

### Author Contributions and Guarantor Statement

P.R.V.O.S and A.Q researched data, contributed to discussion, and wrote the first draft of the manuscript. W.D, S.V.D, K.N, S.R and S.A reviewed and edited the manuscript. P.R.V.O.S is the guarantor of this work and, as such, had full access to all the data in the study and takes responsibility for the integrity of the data and the accuracy of the data analysis.

## Data Availability

All data produced in the present study are available upon reasonable request to the authors.

https://www.cdc.gov/diabetes/data/index.html

https://www.countyhealthrankings.org/explore-health-rankings/rankings-data-documentation

https://www.epa.gov/ejscreen/download-ejscreen-data

## Notes

**<u>Disclosures:</u>** The authors declare no conflicts of interest.

### Competing Interest Statement

The authors have declared no competing interest.

### Funding Statement

This study did not receive any funding

### Author Declarations

The study used (or will use) ONLY openly available human data that were originally located at the Diabetes Surveillance System (DSS) from the Centers for Disease Control (CDC), and County Health Rankings & Roadmaps. Link: https://www.cdc.gov/diabetes/data/index.html link: https://www.countyhealthrankings.org/explore-health-rankings/rankings-data-documentation

## References

1. Blüher M. Obesity: global epidemiology and pathogenesis. Nat Rev Endocrinol. 2019;15(5):288–298. doi:10.1038/s41574-019-0176-8

2. Ward ZJ, Bleich SN, Cradock AL, et al. Projected U.S. State-Level Prevalence of Adult Obesity and Severe Obesity. N Engl J Med. 2019;381(25):2440–2450. doi:10.1056/NEJMsa1909301

3. Lakerveld J, Mackenbach J. The Upstream Determinants of Adult Obesity. Obes Facts. 2017;10(3):216–222. doi:10.1159/000471489

4. Swinburn B, Egger G, Raza F. Dissecting Obesogenic Environments: The Development and Application of a Framework for Identifying and Prioritizing Environmental Interventions for Obesity. Prev Med. 1999;29(6):563–570. doi:10.1006/pmed.1999.0585

5. Mills CW, Johnson G, Huang TTK, Balk D, Wyka K. Use of Small-Area Estimates to Describe County-Level Geographic Variation in Prevalence of Extreme Obesity Among US Adults. JAMA Netw Open. 2020;3(5):e204289. doi:10.1001/jamanetworkopen.2020.4289

6. Ludwig J, Sanbonmatsu L, Gennetian L, et al. Neighborhoods, Obesity, and Diabetes — A Randomized Social Experiment. N Engl J Med. 2011;365(16):1509–1519. doi:10.1056/NEJMsa1103216

7. Tyrrell J, Jones SE, Beaumont R, et al. Height, body mass index, and socioeconomic status: mendelian randomisation study in UK Biobank. The BMJ. 2016;352:i582. doi:10.1136/bmj.i582

8. Racial-Ethnic Disparities in Obesity and Biological, Behavioral, and Sociocultural Influences in the United States: A Systematic Review - PMC. Accessed August 23, 2023. https://www.ncbi.nlm.nih.gov/pmc/articles/PMC8433490/

9. Dong W, Bensken WP, Kim U, et al. Variation in and Factors Associated With US County-Level Cancer Mortality, 2008-2019. JAMA Netw Open. 2022;5(9):e2230925. doi:10.1001/jamanetworkopen.2022.30925

10. Dong W, Bensken WP, Kim U, Rose J, Berger NA, Koroukian SM. Phenotype Discovery and Geographic Disparities of Late-Stage Breast Cancer Diagnosis across U.S. Counties: A Machine Learning Approach. Cancer Epidemiol Biomarkers Prev. 2022;31(1):66–76. doi:10.1158/1055-9965.EPI-21-0838

11. US EPA O. EJScreen: Environmental Justice Screening and Mapping Tool. Published September 3, 2014. Accessed April 5, 2023. https://www.epa.gov/ejscreen

12. Dong W, Motairek I, Nasir K, et al. Risk factors and geographic disparities in premature cardiovascular mortality in US counties: a machine learning approach. Sci Rep. 2023;13:2978. doi:10.1038/s41598-023-30188-9

13. Products - Data Briefs - Number 360 - February 2020. Published June 26, 2020. Accessed August 26, 2023. https://www.cdc.gov/nchs/products/databriefs/db360.htm

14. Wang Y, Beydoun MA, Min J, Xue H, Kaminsky LA, Cheskin LJ. Has the prevalence of overweight, obesity and central obesity levelled off in the United States? Trends, patterns, disparities, and future projections for the obesity epidemic. Int J Epidemiol. 2020;49(3):810–823. doi:10.1093/ije/dyz273

15. Smyth S, Heron A. Diabetes and obesity: the twin epidemics. Nat Med. 2006;12(1):75–80. doi:10.1038/nm0106-75

16. Dietary Guidelines for Americans, 2020-2025.

17. Pollack A. A.M.A. Recognizes Obesity as a Disease. The New York Times. https://www.nytimes.com/2013/06/19/business/ama-recognizes-obesity-as-a-disease.html. xPublished June 18, 2013. Accessed August 26, 2023.

18. Roberto CA, Swinburn B, Hawkes C, et al. Patchy progress on obesity prevention: emerging examples, entrenched barriers, and new thinking. The Lancet. 2015;385(9985):2400–2409. doi:10.1016/S0140-6736(14)61744-X

19. Kyle TK, Dhurandhar EJ, Allison DB. Regarding Obesity as a Disease: Evolving Policies and Their Implications. Endocrinol Metab Clin North Am. 2016;45(3):511–520. doi:10.1016/j.ecl.2016.04.004

20. Silveira EA, Mendonça CR, Delpino FM, et al. Sedentary behavior, physical inactivity, abdominal obesity and obesity in adults and older adults: A systematic review and meta-analysis. Clin Nutr ESPEN. 2022;50:63–73. doi:10.1016/j.clnesp.2022.06.001

21. Tarp J, Rossen J, Ekelund U, Dohrn IM. Joint associations of physical activity and sedentary time with body mass index: A prospective study of mortality risk. Scand J Med Sci Sports. 2023;33(5):693–700. doi:10.1111/sms.14297

22. de Araújo M do ESC, da Conceição Chagas de Almeida M, Matos SMA, de Jesus Mendes da Fonseca M, Pitanga CPS, Pitanga FJG. Combined Effect of Leisure-Time Physical Activity and Sedentary Behavior on Abdominal Obesity in ELSA-Brasil Participants. Int J Environ Res Public Health. 2023;20(15):6501. doi:10.3390/ijerph20156501

23. Mokdad AH, Ford ES, Bowman BA, et al. Prevalence of Obesity, Diabetes, and Obesity-Related Health Risk Factors, 2001. JAMA. 2003;289(1):76–79. doi:10.1001/jama.289.1.76

24. Kirby JB, Liang L, Chen HJ, Wang Y. Race, Place, and Obesity: The Complex Relationships Among Community Racial/Ethnic Composition, Individual Race/Ethnicity, and Obesity in the United States. Am J Public Health. 2012;102(8):1572–1578. doi:10.2105/AJPH.2011.300452

25. Woolcott OO, Seuring T. Temporal trends in obesity defined by the relative fat mass (RFM) index among adults in the United States from 1999 to 2020: a population-based study. BMJ Open. 2023;13(8):e071295. doi:10.1136/bmjopen-2022-071295

26. Elia M. Obesity in the Elderly. Obes Res. 2001;9(S11):244S–248S. doi:10.1038/oby.2001.126

27. Schwartz MW, Seeley RJ, Zeltser LM, et al. Obesity Pathogenesis: An Endocrine Society Scientific Statement. Endocr Rev. 2017;38(4):267–296. doi:10.1210/er.2017-00111

28. Darmon N, Drewnowski A. Does social class predict diet quality? Am J Clin Nutr. 2008;87(5):1107–1117. doi:10.1093/ajcn/87.5.1107

29. Hanson KL, Connor LM. Food insecurity and dietary quality in US adults and children: a systematic review123. Am J Clin Nutr. 2014;100(2):684–692. doi:10.3945/ajcn.114.084525

30. Crawford PB, Webb KL. Unraveling the Paradox of Concurrent Food Insecurity and Obesity. Am J Prev Med. 2011;40(2):274–275. doi:10.1016/j.amepre.2010.11.003

31. Hohl A, Lotfata A. Modeling spatiotemporal associations of obesity prevalence with biking, housing cost and green spaces in Chicago, IL, USA, 2015–2017. J Transp Health. 2022;26:101412. doi:10.1016/j.jth.2022.101412

32. Lotfata A, Tomal M. Exploring Housing Determinants of Obesity Prevalence Using Multiscale Geographically Weighted Regression in Chicago, Illinois. Prof Geogr. 2023;75(3):335–344. doi:10.1080/00330124.2022.2111692

33. Brakefield WS, Olusanya OA, Shaban-Nejad A. Association Between Neighborhood Factors and Adult Obesity in Shelby County, Tennessee: Geospatial Machine Learning Approach. JMIR Public Health Surveill. 2022;8(8):e37039. doi:10.2196/37039

34. Scheinker D, Valencia A, Rodriguez F. Identification of Factors Associated With Variation in US County-Level Obesity Prevalence Rates Using Epidemiologic vs Machine Learning Models. JAMA Netw Open. 2019;2(4):e192884. doi:10.1001/jamanetworkopen.2019.2884

35. Ayanian JZ, Weissman JS, Schneider EC, Ginsburg JA, Zaslavsky AM. Unmet Health Needs of Uninsured Adults in the United States. JAMA. 2000;284(16):2061–2069. doi:10.1001/jama.284.16.2061

36. Collazo-Clavell ML. Managing Obesity: Scaling the Pyramid to Success. Mayo Clin Proc. 2019;94(6):933–935. doi:10.1016/j.mayocp.2019.04.011

37. Recommended Community Strategies and Measurements to Prevent Obesity in the United States. Accessed August 31, 2023. https://www.cdc.gov/mmwr/preview/mmwrhtml/rr5807a1.htm

38. Avenue 677 Huntington, Boston, Ma 02115. Measuring Obesity. Obesity Prevention Source. Published October 21, 2012. Accessed August 30, 2023. https://www.hsph.harvard.edu/obesity-prevention-source/obesity-definition/how-to-measure-body-fatness/

39. Dong W, Kim U, Rose J, et al. Geographic Variation and Risk Factor Association of Early Versus Late Onset Colorectal Cancer. Cancers. 2023;15(4):1006. doi:10.3390/cancers15041006

40. THE MAX□JP□JREGIONS PROBLEM* - Duque - 2012 - Journal of Regional Science - Wiley Online Library. Published May 15, 2023. Accessed May 15, 2023. https://onlinelibrary.wiley.com/doi/full/10.1111/j.1467-9787.2011.00743.x?saml_referrer

