## Supplemental Figure 1, Supplemental Table 1 for "County-level Socio-Environmental Factors and Obesity Prevalence in the United States"

**Supplemental Figure 1.** Median county-level obesity prevalence percentages by county socio-enviromic cluster as predicted by CART. No statistical difference between training and testing set in relation to cluster mortality was observed. Training set (n=2204 US counties), Test set (n=548 US counties).

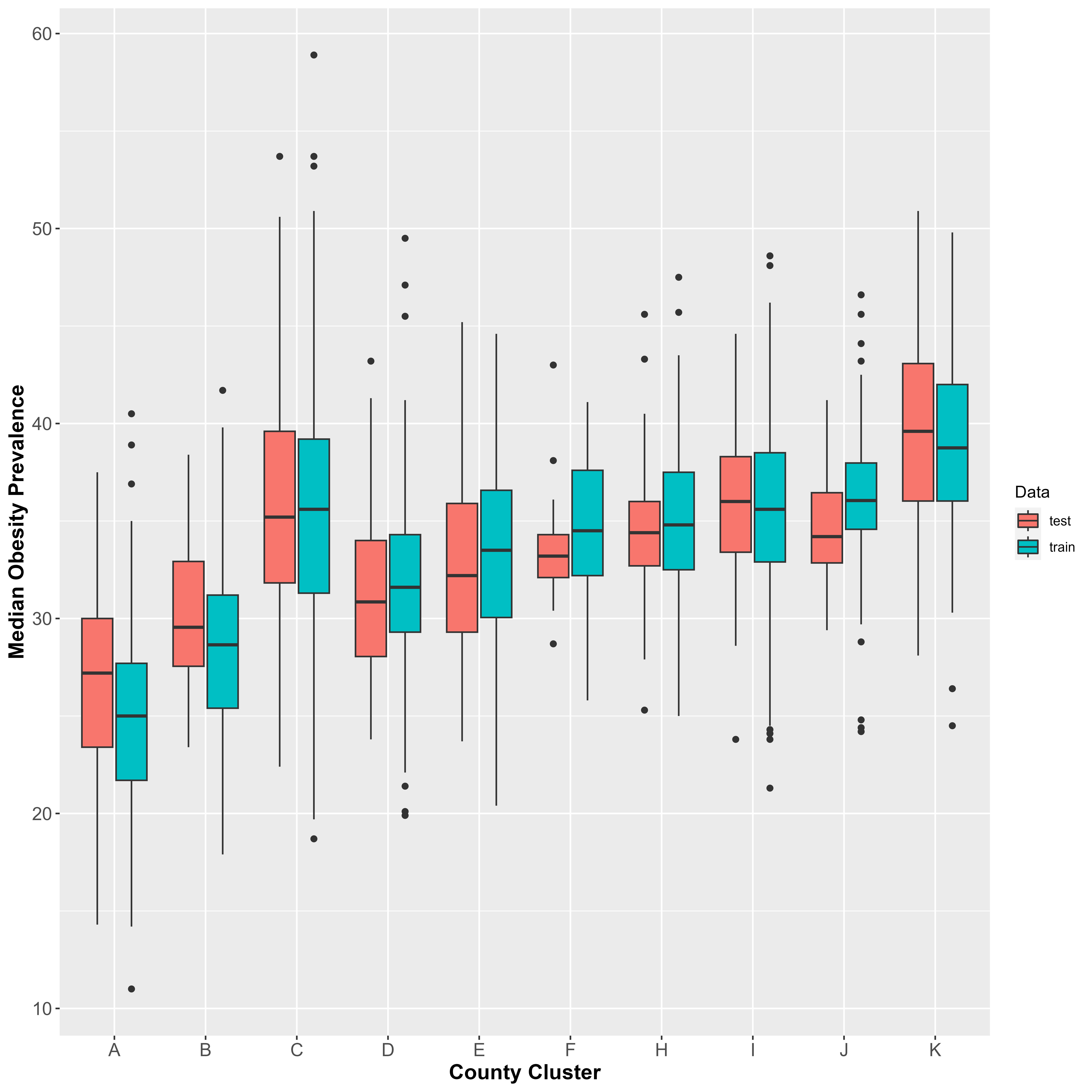

**Supplemental Table 1.** Definition and source of variables included in this study.

| Variable | Description | Year | Original Source |
| --- | --- | --- | --- |
| **Race/Ethnicity and Population Structure** | | | |
| Hispanic Population | Percentage of population identifying as Hispanics | 2017 | Census -PE |
| Non-Hispanic White | Percentage of Non-Hispanic White population | 2017 | Census -PE |
| Non-Hispanic Black | Percentage of Non-Hispanic African American population | 2017 | Census -PE |
| Asian and Pacific Islander | Percentage of Asian and Pacific Islander population | 2017 | Census -PE |
| Female Population | Percentage of female population | 2017 | Census -PE |
| Rural Population | Percentage of people living in rural areas | 2010 | Census -PE |
| Population above 65 years | Percentage of population age 65 years or older | 2017 | Census -PE |
| Population under 18 years | Percentage of population age 18 years or younger | 2017 | Census -PE |
| **Environmental Exposure** | | | |
| Particulate Matter 2.5 Level in Air | PM­_2.5_ levels in air, µg/m3 (annual average) | 2018 | EPA - EJSCREEN |
| Diesel Particulate Matter Level in Air | Diesel particulate matter level in air, µg/m3 | 2017 | EPA- EJSCREEN |
| Ozone Level in Air | Ozone summer seasonal average of daily maximum 8 h concentration in air in parts per billion | 2018 | EPA- EJSCREEN |
| Air Toxics Cancer Risk | Lifetime exposure to air toxics cancer risk (in persons per million) | 2017 | EPA- EJSCREEN |
| Proximity to NPL Sites | Count of proposed or listed NPL—also known as superfund—sites within 5 km (or nearest one beyond 5 km), each divided by distance in km | 2020 | EPA - EJSCREEN |
| Proximity to RMP Sites | Count of RMP (potential chemical accident management plan) facilities within 5 km (or nearest one beyond 5 km), each divided by distance in km | 2020 | EPA - EJSCREEN |
| Traffic Proximity and Volume | Average annual daily count of vehicles at major roads within 500 m, divided by distance in meters | 2019 | EPA - EJSCREEN |
| **Health Behaviors and Conditions** | | | |
| Adult Smoking | Percentage of adults who are current smokers (age-adjusted) | 2017 | CDC - BRFSS |
| Obesity | Percentage of the adult population (aged ≥18 y) that reports a body mass index ≥30 (age-adjusted) | 2017 | CDC - DSS |
| Diabetes | Percentage of adults (age ≥ 20) with diagnosed diabetes (age-adjusted) | 2017 | CDC - DSS |
| Low Birthweight | Percentage of live births with low birthweight (< 2,500 grams) | 2013-2019 | CDC - NCHS |
| Flu Vaccinations | Percentage of fee-for-service Medicare enrollees that had an annual flu vaccination | 2017 | CMS -MMD |
| Excessive Drinking | Percentage of adults reporting binge or heavy drinking (age-adjusted) | 2017 | CDC - BRFSS |
| Access to Exercise Opportunities | Percentage of people with adequate access to locations for physical activity | 2010, 2019 | ESRI & Census -TF |
| Physically Inactive | Percentage of adults (age ≥ 18 y) reporting no leisure-time physical activity (age-adjusted) | 2017 | CDC- DSS |
| Poor Mental Health Days | Average number of mentally unhealthy days reported in past 30 days (age-adjusted). | 2019 | CDC - BRFSS |
| Insufficient Sleep | Percentage of adults who report fewer than 7 hours of sleep on average (age-adjusted). | 2018 | CDC - BRFSS |
| Poor Physical Health Days | Average number of physically unhealthy days reported in past 30 days (age-adjusted). | 2019 | CDC - BRFSS |
| Frequent Mental Distress | Percentage of adults reporting 14 or more days of poor mental health per month (age-adjusted). | 2019 | CDC - BRFSS |
| Frequent Physical Distress | Percentage of adults reporting 14 or more days of poor physical health per month (age-adjusted). | 2019 | CDC - BRFSS |
| **Socioeconomic Factors** | | | |
| Median Household Income | The income (US dollar) where half of households in a county earn more and half of households earn less | 2017 | AHRF |
| Unemployment | Percentage of people (aged≥16 year) unemployed but seeking work | 2017 | BLS |
| Income Inequality | Ratio of household income at the 80th percentile to income at the 20th percentile | 2015-2019 | Census - ACS |
| Poverty | Percentage of people whose income under the federal poverty level | 2017 | AHRF |
| Under 200% Poverty (18-64 years) | Percentage of people (aged 18–64 year) whose income is under 200% of the federal poverty level | 2017 | AHRF |
| Children in Poverty | Percentage of people under age 18 in poverty. | 2020 | Census - SAIPE |
| High School Completion | Percentage of people aged ≥ 25 years with a high school diploma or equivalent | 2015-2019 | Census - ACS |
| Some College Degree | Percentage of people (aged 25–44 year) with some post-secondary education | 2015-2019 | Census - ACS |
| Severe Housing Problems | Percentage of households with at least 1 of 4 housing problems: overcrowding, high housing costs, lack of kitchen facilities, or lack of plumbing facilities | 2013-2017 | CHAS |
| Severe Housing Cost Burden | Percentage of households that spend 50% or more of their household income on housing | 2015-2019 | Census-ACS |
| Homeownership | Percentage of owner-occupied housing units | 2015-2019 | Census - ACS |
| Children in Single-Parent Household | Percentage of children that live in a household headed by a single parent. | 2016-2020 | Census - ACS |
| Limited Access to Healthy Food | Percentage of people who are low-income and do not live close to a grocery store | 2015 | USDA -FEA |
| Food Stamp Recipients | Percentage of people who were food stamp recipients | 2017 | AHRF |
| Food Insecurity | Percentage of people who lack adequate access to food | 2017 | MMG |
| Food Environment Index | Index of factors that contribute to a healthy food environment, from 0 (worst) to 10 (best). | 2019 | USDA - FEA |
| Social Association | Number of membership associations per 10,000 population | 2017 | CBP |
| Injury Deaths | Number of deaths due to injury per 100,000 population. | 2016-2020 | NVSS |
| Uninsured Rate | Percentage of people (aged 18–64 year) without health insurance | 2017 | HRSA - AHRF |
| Long Commute – Driving Alone | Among workers who commute in their car alone, the percentage that commute more than 30 minutes. | 2016-2020 | Census -ACS |
| Primary Care Physicians | Primary care physicians in patient care per 100,000 people | 2017 | HRSA -AHRF |
| Broadband Access | Percentage of households with broadband internet connection | 2015-2019 | Census -ACS |

Abbreviations: AHRF: Area Health Resources Files; ACS: American Community Survey; BLS: Bureau of Labor Statistics; BRFSS: Behavioral Risk Factor Surveillance System; CBP: County Business Patterns; CDC: Centers for Disease Control and Prevention; CHAS: Comprehensive Housing Affordability Strategy; CHF: County Health Rankings & Roadmaps; CMS: Centers for Medicare & Medicaid Services; DSS: US Diabetes Surveillance System; EJSCREEN: Environmental Justice Screening tool ; EPA: Environmental Protection Agency; FEA: Food Environment Atlas; HRSA: Health Resources and Services Administration; MMD: Mapping Medicare Disparities (MMD) Tool; MMG: Map the Meal Gap; NCHHSTP: National Center for HIV/AIDS, Viral Hepatitis, STD, and TB Prevention; NCHS: National Center for Health Statistics; NLP: National Priorities List; PE: Population Estimates; PM: Fine particulate matter; RMP: Risk Management Plan; USDA: US Department of Agriculture.
